# Online General Exercise Management (GEM) Program for patients with Breast Cancer-Related Lymphedema: A Protocol for a Pilot Randomized Controlled Trial

**DOI:** 10.1101/2025.01.20.25320868

**Authors:** Yoko Arinaga, Fumiko Sato, Satoshi Miyata, Neil Piller, Tohru Ohtake

## Abstract

**Background:** Breast cancer-related lymphedema (BCRL) requires a lifetime commitment to self-care.

**Aims:** This is a study protocol for a pilot parallel-group randomized controlled trial which will assess the effectiveness of an online version of the General Exercise Management (GEM) program for patients with BCRL.

**Methods:** Sixty patients will be recruited through online advertisements and randomly allocated to either an intervention or control group. The intervention group will be provided with an educational program on standard self-care for BCRL as well as the GEM program, while the control group will receive only the self-care education program. Both groups will participate in a six-month online program, perform daily self-checks and participate in monthly small-group sessions. All participants, session facilitators and a statistical analyst will be blinded to group allocation.

**Anticipated Results:** The primary expected outcome is a reduction of BCRL symptoms, and the secondary outcomes are health-related quality of life and self-care adherence improvements.

**Conclusions:** The findings will inform further investigation in a Phase III trial.

**Declaration of Interest:** All authors declare that they have no competing financial interests.

This study was approved by the Institutional Review Board of Fukushima Medical University Hospital (REC2024-068) and registered with UMIN-CTR (UMIN000055357).

## Background

Breast cancer-related lymphedema (BCRL) is a chronic condition affecting approximately 29.8% of breast cancer survivors (Shen et al., 2024). BCRL symptoms typically include arm swelling, pain, and various physical discomforts that affect daily activities(Shen et al., 2024). These symptoms significantly impact a patient’s long-term quality of life (QOL) (Ramirez-Parada et al., 2023). Managing BCRL requires a lifetime commitment to self-care, but many patients find it challenging to maintain such a commitment (Fu et al., 2023).

Complete decongestive therapy (CDT) is the most commonly recommended approach for improving health-related quality of life (HRQOL) and managing lymphedema (Rangon et al., 2022, JLS, 2024). CDT consists of skin care, exercise, compression therapy, and daily self-care education, including weight management. Exercise has been shown to have benefits for BCRL patients (Lin et al., 2023, Ali et al., 2021, Hasenoehrl et al., 2020) and the clinical practice guidelines published by the Japanese Lymphedema Society in 2024 (JLS, 2024) also recommend exercise as part of the treatment plan for BCRL. In this guideline (JLS, 2024), several studies of various exercises are included (Wang et al., 2023, Kilbreath et al., 2020, Pirincci et al., 2023, Pasyar et al., 2019). Despite the importance of exercise, existing Japanese guidelines often lack specific recommendations regarding the type, frequency, and duration of exercise, making it difficult for healthcare providers to instruct patients on specific methods.

Although high-frequency exercise programs have been reported to reduce the severity of symptoms (Lian et al., 2024), only 14.2% of lymphedema treatment facilities in Japan implement exercise therapy (Ishii et al., 2021), which may explain poor patient adherence. Furthermore, the complexity of self-care, as well as the various costs associated with practicing it, place a considerable burden on both patients and caregivers, often resulting in low adherence and worsening of the conditions (Lytvyn et al., 2020). Patients must be provided with education on practical and feasible self-care in order to encourage adherence.

We (Arinaga et al., 2019) previously reported the benefits of a 10-minute holistic self-care program for BCRL patients, which included: a modified Radio Taiso No.1 (a calisthenics program that has long been practiced in Japan) (Japan-Post- Insurance); gentle arm exercises with deep breathing (Moseley et al., 2005), central lymphatic drainage; and skin care using modified traditional lymphatic drainage techniques(Arinaga et al., 2019).

The control group continued with the usual care as recommended by their respective hospitals over a 6-month period. In our program, Radio Taiso was modified to be performed at half the speed of the original for BCRL patients, in order to prevent damage to contracted muscles or surgical scars, reduce peripheral movement of lymphatic fluid caused by centrifugal force and enhance muscle strength by maintaining muscle tension for a longer duration.

The intervention group showed significant reductions in relative hand edema volume and improvements in most BCRL symptoms compared to the control group, who showed improvements only in pain and coldness. While the program was beneficial, the effects of its individual components were not evaluated separately. Online interventions have shown benefits in lymphedema management and patient recruitment (Brøgger-Mikkelsen et al., 2020).

## Aims

This study aims to evaluate the efficacy of an Online General Exercise Management (GEM) based on modified Radio Taiso, plus standard self-care education compared to standard self-care education only, focusing on BCRL symptoms, health-related quality of life, and self-care adherence.

## Methods

### Participants

Sixty participants (30 for each group) in Japan, who will have self-confirmed their eligibility as detailed in the following criteria, will be recruited using Online advertisements. (figure 1)

### Inclusion Criteria

*Adults aged 18 years or older

*History of breast cancer surgery

*Persistent BCRL symptoms (e.g., swelling of the upper extremity on the surgery side) for at least 3 months post-treatment

*Ability to understand study materials and complete online questionnaires

*Access to the internet and ability to attend monthly online sessions

### Exclusion Criteria

*Current recurrence of breast cancer or presence of other cancers

*History of radiotherapy or chemotherapy within the last 6 months, or plans to undergo either during the study period

*Current infection, inflammation, or deep vein thrombosis in the arm on the surgery side

*Difficulty responding to online questionnaires

### Timeline

The study will be conducted from August 2024 to March 2028, including enrollment and follow-up periods.

### Randomization and Blinding

Participants will be randomly assigned in a 1:1 ratio to either the intervention group or control group using block randomization with computer-generated random numbers. In this study, participants, small-group session facilitators, and statistical analysts will be blinded to group assignments to minimize bias.

### Interventions

Both groups will participate in a 6-month online program featuring standard self-care education based on the recent guidelines (JLS, 2024) and research, including videos and texts on BCRL knowledge, skin care, exercise, compression therapy, weight management, and other self-care practices. All participants will complete daily self-care checklists and attend monthly online small-group sessions facilitated by trained staff to share experiences and receive support.

### Control Group

Participants will have access to the standard self-care education program website only.

### Intervention Group

In addition to the above standard self-care education, the intervention group will receive access to the GEM program. This program is based on Radio Taiso No. 1 (Japan-Post-Insurance) and was adapted for BCRL patients by reducing movement speed by half for therapeutic purposes (Arinaga et al., 2019).

### Primary Outcome to be assessed

This is the rate of change in the total BCRL symptom score from baseline to 6 months. The symptoms of BCRL will be assessed using an online self-completed questionnaire consisting of 11 items that were selected based on previous literature (Keeley et al., 2010, Launois et al., 2002). The questionnaire has been validated for face validity by breast surgeons, dermatologists, and breast cancer nursing researchers (Arinaga et al., 2019). Symptoms in the affected upper limb will be self-reported using a seven-point semantic differential scale, ranging from 0 (none) to 6 (very severe). The total score will range from 0 to 72, with higher scores indicating a greater severity and number of symptoms.

### Secondary Outcomes to be assessed

Secondary outcomes will be evaluated at baseline, 1 week, 1 month, 3 months, and 6 months using an online questionnaire.

This will include:

1. Individual BCRL symptom scores
2. Health-related quality of life using the validated Japanese SF-8 (Fukuhara and Suzukamo, 2014) instrument.
3. Self-care adherence measured by daily performance of skin moisturizing (three times daily), exercise (twice daily for intervention group), weight monitoring, and compression therapy (when prescribed), with scores: 0 (not performed), 1 (performed 1-2 times/week), 2 (performed 3-5 times/week), and 3 (performed almost every day)
4. Self-perceived effectiveness and burden of self-care rated on a 7-point semantic differential scale ranging from 0 (none) to 6 (very much)
5. Daily time spent on self-care activities
6. Adverse events

### Sample Size Calculation

Sample size calculation is based on previous pilot studies (Basha et al., 2022, Whitehead et al., 2016). Using an effect size of 0.4, 80% power, and a two-sided 5% alpha level, we determined that 60 participants (30 per group) would be needed, including a 20% allowance for potential dropout.

### Data Collection and Analysis

#### Primary Outcome Analysis

The primary outcome will be analyzed using the FAS. The analysis will focus on the change in total BCRL symptom scores from baseline to 6 months. The difference in mean scores between the intervention and control groups will be tested using Welch’s t-test or the Wilcoxon rank-sum test, as appropriate. A *p*- value of < 0.05 will be considered statistically significant.

#### Secondary Outcome Analysis

For continuous variables, group comparisons will be performed at each time point (baseline, 1 week, 1 month, 3 months, and 6 months) using Welch’s t-test and the Wilcoxon rank-sum test. For categorical variables, Fisher’s exact test will be used for group comparisons. To address multiple comparisons, adjustments will be made using Holm’s method.

For longitudinal data, multivariate analyses will be conducted using a linear mixed-effects model.

#### Additional Analyses

Summary statistics will be calculated for all variables, with continuous data presented as mean (SD) and categorical data as frequencies (%). Adverse events will be documented and compared between groups. Sensitivity analyses will include PPS analysis and multiple imputation for missing data. Outliers will be included in the primary analysis, with sensitivity analyses conducted if necessary.

## Discussion

This Phase II trial aims to evaluate the Online General Exercise Management (GEM) Program for BCRL patients, based on modified Radio Taiso, a widely recognized Japanese calisthenics program. The GEM program’s cultural familiarity and simplicity may enhance self-care adherence. Additionally, the online delivery of the intervention addresses the accessibility challenges often faced by patients with lymphedema, particularly those in rural or underserved areas.

The results of this study will provide foundational data for the planning of a Phase III trial. Furthermore, the online format offers a cost-effective solution that can be easily expanded to reach more patients, addressing the challenges of in-person care, particularly during global health crises such as the COVID-19 pandemic.

Potential limitations of this study include the reliance on self-reported outcomes and variability in participants’ engagement with the online platform. Additionally, the use of online recruitment may introduce selection bias, as only participants with internet access will be eligible. Future studies should incorporate a more objective method to assess changes in lymphedema symptoms and increase the sample size. It will also be essential to disseminate the effectiveness of the GEM program to a broader range of BCRL patients—who must maintain lifelong self-care for lymphedema—and to the healthcare providers who support them, as the program is designed to be simple and suitable for long-term implementation.

## Data Availability

All data produced in the present study are available upon reasonable request to the authors

## Acknowledgements

This work was supported by JSPS KAKENHI Grant Number 23K09964. We would like to express our sincere gratitude to Professor Takeyasu Kakamu at Fukushima Medical University for his valuable statistical advice. We employed AI-based tools (ChatGPT and Claude 3.5 Sonnet) to assist with summarizing literature reviews and producing paraphrases. The accuracy and integrity of the final manuscript has been reviewed and confirmed by all authors and proofread by native English-speaking scientific editors.

